# Can adolescent depressive symptoms act as a target for reducing intergenerational smoking transmission?

**DOI:** 10.64898/2025.12.04.25341611

**Authors:** Alexandria Andrayas, Elinor Curnow, Jasmine Khouja, Jon Heron, Lindsey Hines, Hannah Jones, Marcus Munafò, Gemma Hammerton, Hannah Sallis

## Abstract

**Background:** Parental smoking increases offspring smoking risk. Young people with mental health difficulties are more likely to have parents who smoke and start smoking themselves. Improving mental health in youth could help disrupt intergenerational cycles of nicotine use, but the extent to which depressive symptoms mediate the link between parental and offspring smoking remains unclear.

**Methods:** We used data on 6,729 participants of the Avon Longitudinal Study of Parents and Children (ALSPAC) with mother-reported data on parental smoking. Counterfactual-based mediation analysis was used to estimate both total and direct effects of parental smoking at offspring age 12 on offspring smoking at age 16 and estimate the indirect effect via offspring depressive symptoms at age 14. We also accounted for early offspring substance use (cigarettes, alcohol, or cannabis) at age 13. Multivariable logistic regression was used to examine pairwise associations between these variables.

**Results:** Mediation analysis showed a total effect of parental smoking on offspring smoking (odds ratio [OR] = 1.44, 95% CI = 1.20-1.72). There was no indirect effect via depressive symptoms (OR = 1.00, 95% CI = 0.99-1.01) when adjusted for confounding. Regression analyses showed parental smoking was associated with early offspring substance use and later offspring smoking but not offspring depression. Early offspring substance use was associated with both offspring depression and later smoking. Higher depressive symptoms were associated with later smoking.

**Conclusions:** Parental smoking in early adolescence increases the odds of offspring smoking in later adolescence, but this does not appear to act through offspring depression.

## Introduction

Children are known to acquire behaviours by observing and imitating role-models such as parents (Bandura & Walters, 1977) and this has been shown experimentally (Burstein & Ginsburg, 2010; Shimpi, Akhtar, & Moore, 2013). The influence of parental smoking on offspring smoking behaviours is one example of this with highly detrimental implications. Studies consistently show that children of people who smoke are more likely to start smoking themselves than children of non-smokers (Leonardi-Bee, Jere, & Britton, 2011; Lochbuehler, Schuck, Otten, Ringlever, & Hiemstra, 2016; Mays et al., 2014; Vitória, Pereira, Muinos, Vries, & Lima, 2020), acting through mechanisms such as modelling of smoking behaviour (White, Johnson, & Buyske, 2000) and altered social norms within the household (Dickter, Forestell, & Volz, 2018). This perpetuates cycles of nicotine dependence across generations (Alves et al., 2017; Gray & Squeglia, 2018).

Parental smoking is also associated with a range of offspring outcomes that extend beyond health behaviours, including cognitive, neurodevelopmental, mental health and behavioural outcomes (Easey & Sharp, 2021; Rice, Langley, Woodford, Smith, & Thapar, 2018; Thapar & Rutter, 2009). However there is uncertainty in this evidence where some of these complex associations may reflect causal pathways, and others may arise from shared familial, social, or genetic vulnerabilities (Eilertsen et al., 2021; Gage, Bowden, Davey Smith, & Munafò, 2018; Harrison, Munafò, Davey Smith, & Wootton, 2020; Hicks et al., 2021; Khouja, Wootton, Taylor, Smith, & Munafò, 2021; Liu et al., 2019; Schellhas et al., 2021; Taylor et al., 2014; Thapar & Rutter, 2009).

Much of the literature has looked at biological mechanisms to explain associations between parental smoking and offspring outcomes. These include *in utero* exposure (Duko, Ayano, Pereira, Betts, & Alati, 2020), shared genetic liability (Jami, Hammerschlag, Bartels, & Middeldorp, 2021), and exposure to second-hand tobacco smoke (Patten et al., 2018; van der Eijk & Woh, 2023). However, children of parents who smoke may also experience poorer mental health through other family and environmental processes, including stigma surrounding parental smoking (Burgess, Fu, & van Ryn, 2009; Stuber, Galea, & Link, 2008), economic strain (Lakdawala & Simon, 2017), parental ill health (Morris, Turnbull, Preen, Zajac, & Martini, 2018), or disruptions to the parent-child relationship (Brook, Balka, Fei, & Whiteman, 2006; Woodgate & Kreklewetz, 2012). These mechanisms illustrate why offspring mental health may plausibly be implicated in the link between parental smoking and smoking in offspring.

Young people with mental health issues such as depression and anxiety, lower cognitive abilities, and behavioural disorders are themselves more prone to start smoking (Chaiton, Cohen, O’Loughlin, & Rehm, 2009; Daly & Egan, 2017; Fluharty, Taylor, Grabski, & Munafò, 2017; Hockenberry, Timmons, & Weg, 2011). Additional evidence suggests the combination of parental smoking and lower offspring mental health competence (i.e., worse prosocial behaviours and learning skills) has an additional interactive effect on offspring smoking (Pearce et al., 2021). Addressing psychological health in smoking cessation efforts has also been shown to be effective (Lightfoot, Panagiotaki, & Nobes, 2020). Taken together, this suggests that improving offspring mental health could help reduce their likelihood of smoking. What is less clear is the extent to which offspring mental health contributes to the intergenerational transmission of smoking as opposed to reflecting shared underlying factors.

Traditional observational studies face challenges in determining whether intergenerational associations are causal, owing to residual confounding and shared familial influences that are difficult to separate (Lawlor, Davey Smith, Kundu, Bruckdorfer, & Ebrahim, 2004; Pearce & Lawlor, 2016). These issues remain relevant here. Research on parental smoking predominantly focuses simply on whether their adolescent offspring have ever smoked.

There is a need for comprehensive approaches that examine the long-term impacts of parental smoking, the interplay with offspring mental health, and in turn more fine-grained measures of smoking habits (Balbuena, Mela, & Ahmed, 2024; Jami et al., 2021; Rajyaguru, Kwong, Braithwaite, & Pearson, 2021). This highlights the importance of longitudinal data and causal inference methodologies.

In this study, we make use of rich multigenerational birth cohort data and counterfactual-based mediation analysis (Daniel, De Stavola, & Cousens, 2011; Pearl, 2022) to bridge this gap. We aim to: **a.** explore the associations between parental smoking (offspring age 12), early offspring substance use (age 13), offspring depressive symptoms (age 14) and later offspring smoking (age 16); **b.** investigate the total causal effect of parental smoking on offspring smoking; and **c.** decompose the total effect to examine the extent to which offspring depressive symptoms mediates intergenerational smoking. Better understanding of these relationships will inform better preventive measures and tailored interventions, ultimately reducing the intergenerational transmission of smoking and its related health consequences (Fitzsimons, Goodman, Kelly, & Smith, 2017; Malanchini, Rimfeld, Allegrini, Ritchie, & Plomin, 2020; Prakash & Kumar, 2021).

## Methods Participants

Pregnant women resident in Avon, UK with expected delivery dates between 1st April 1991 and 31st December 1992 were invited to take part in the Avon Longitudinal Study of Parents and Children (ALSPAC). Initially 14,541 pregnancies were enrolled, and 13,988 children were alive at 1 year of age. When the oldest children were approximately 7 years of age, an attempt to bolster the initial sample meant the total sample size for analyses using any data collected thereafter is 15,447 pregnancies, where 14,901 children were alive at 1 year of age. 14,203 unique mothers initially enrolled, but due to additional phases of recruitment, 14,833 unique women were enrolled in ALSPAC as of September 2021.

12,113 partners of these mothers have been in contact with the study and 3,807 are currently enrolled (Boyd et al., 2013; Fraser et al., 2013; K. Northstone et al., 2023; Northstone et al., 2019).

The sample used in the present analyses (n = 6,729) consists of offspring participants in ALSPAC who have available parental smoking data at offspring age 12 years. This is referred to as the “analytic sample” throughout.

## Measures

### Parental smoking at offspring age 12 years

Parental smoking, measured when offspring were approximately 12 years of age (via questionnaires sent out in 2007), was defined using maternal reports. The participant’s mother was asked to complete questions asking separately how many cigarettes she and her partner (if applicable) smoked on a weekday or weekend day at the time of the questionnaire. Participants were classified as having a parent who smoked if either their mother or their mother’s partner, or both, smoked at least one cigarette per day. If the mother reported not having a partner, this was based solely on her smoking habits.

### Early offspring substance use at 13 years

Information on offspring substance use at approximately 13 years of age was collected during an interview conducted at an in-person clinic. Interviewers asked whether they had used any substances and the nature of their use. Here a binary measure was derived based on whether the participant stated that they had smoked cigarettes, used cannabis, or drank alcohol without parent’s permission in the past 6 months. Participants who had used one or more substances in this time frame were classified as exhibiting early substance use.

### Offspring depressive symptoms at 14 years

Information on offspring depressive symptoms at approximately 14 years of age was collected during a further in-person clinic. A computer task was carried out where they were asked to answer 13 questions from the Short Mood and Feelings Questionnaire (SMFQ; Angold, Costello, Messer, & Pickles, 1995). From this, a total score was constructed which sums the responses such that a higher score indicates a lower mood. Here a binary measure was used where participants with a total score of ≥8 were defined as having high depressive symptoms. This cut-off has been shown to have good discrimination for depression in children and adolescents (Angold et al., 1995) and may be preferred over the commonly used cut point of ≥12 when favouring sensitivity over specificity (Eyre et al., 2021).

### Offspring current smoking at 16 years

Information on offspring smoking was collected at approximately 16 years of age via a postal questionnaire sent to the participant. A binary measure of current smoking (*No [reference category (ref)] vs Yes*) was derived based on whether they reported smoking cigarettes either sometimes, weekly, or every day, at the time of the questionnaire.

Participants who reported only smoking once or twice ever, or that they used to but did not at the time, were classed as not smoking.

### Confounders

Analyses were adjusted for many confounders including sex assigned at birth (*Male vs Female*), ethnicity (*White vs ethnically minoritised*), maternal smoking during pregnancy (*No vs Yes*), and the following, also assessed during pregnancy: oldest parent’s age, lowest parental social class (*I/II vs III, IV/V*), lowest parental education (*A level or above vs O level, Vocational/Certificate of Secondary Education*), and neighbourhood quality index. Analyses were also adjusted for Townsend deprivation score quintiles (*Q1 vs Q2-5*), household income per week (*£400+ vs <£400*), highest parental Edinburgh Postnatal Depression Scale (EPDS) score, and parental mental health conditions (*No vs Yes*) at offspring age 8, physical or emotional abuse (*No vs Yes*) at age 9, housing tenure (*Owned/Mortgaged vs Rented/Other*) at offspring age 10, and peer substance use (*No vs Yes*), parental monitoring (*Always/Most of the time vs Never/Hardly ever/Sometimes*), and bullying (*No vs Yes*) at age 13. Early-life confounders were assumed stable, and remaining variables were drawn from the timepoints closest to the exposure. Further details on these measures can be found in Supplementary Text S1 alongside information on how confounding relates to the analyses.

Early offspring substance use at age 13 was included as an intermediate confounder in mediation models as this may be a consequence of parental smoking that could confound the relationship between offspring depressive symptoms and later offspring smoking.

## Statistical analysis

Data curation and mediation analyses using the g-computation formula were carried out in Stata (version 18) (StataCorp, 2023), and all other data processing, statistical analyses and visualisation were carried out in R (version 4.5.1) (R Core Team, 2023). Figure 1 shows the proposed model being investigated.

**Figure 1.**
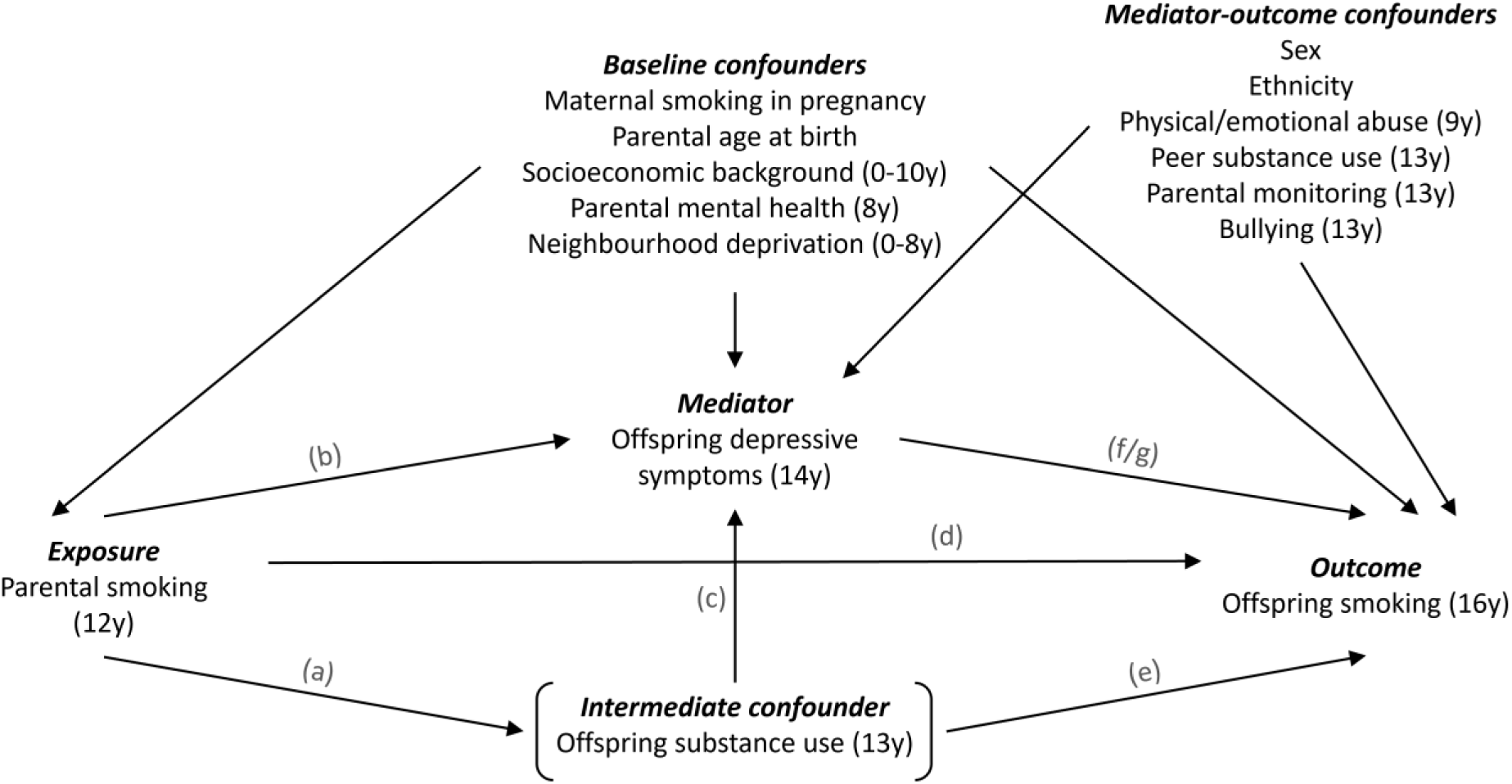
Proposed model. Causal relationships between exposure, mediator, confounders, and outcome are depicted. Lines indicate related variables, with arrows indicating the direction of the relationship; absent lines represent variables with no direct causal relation. Offspring age at the time of measurement is indicated in brackets for each variable. Models fitted: (a) parental smoking at offspring age 12 → early offspring substance use at age 13; (b) parental smoking → offspring depressive symptoms at age 14; (c) early substance use → depressive symptoms (adjusted for parental smoking); (d) parental smoking → offspring current smoking at age 16; (e) early substance use → current smoking (adjusted for parental smoking); (f) depressive symptoms → current smoking (adjusted for parental smoking); (g) depressive symptoms → current smoking (adjusted for parental smoking + early substance use).

### Missing data approach

Patterns of missingness in the analytic sample were investigated. In addition, univariable logistic regression models were used to explore the association between the analysis variables and (i) missingness (i.e., whether all data are reported or not) and (ii) being in the analytic sample. We also looked at (iii) a multivariable logistic regression to explore whether the association between offspring smoking and missingness is evident after adjusting for the parental smoking, offspring depression and a subset of confounders.

Multiple imputation (MI) by chained equations was used to impute missing data up to the analytic sample. We imputed 50 datasets with 25 iterations using the mice R package (version 3.18.0). The imputation predictor matrix is shown in Supplementary Figure S1. As our analysis models include an exposure-mediator interaction, we accounted for this in the imputation models (Tilling, Williamson, Spratt, Sterne, & Carpenter, 2016). Additional measures available at earlier, more complete, waves of data collection were included as auxiliary variables. These are listed in Supplementary Text S2 alongside further information on our imputation strategy. We examined Monte Carlo errors related to each regression model to check if this number of imputations was appropriate.

The mean of the log odds ratio (OR) was computed, and standard errors (SE) were calculated using Rubin’s rules (Rubin, 1987). These pooled parameters are reported throughout. Complete records analyses were also carried out and any substantial differences highlighted.

### Mediation analysis

Counterfactual-based mediation analysis via the parametric g-computation formula by Monte Carlo simulation was used to allow for a causal interpretation of our research question (Daniel et al., 2011; De Stavola, Daniel, Ploubidis, & Micali, 2015). Here this estimates the total causal, pure natural and controlled direct effects of parental smoking on offspring smoking, and the total natural indirect effect via offspring depression. These effects are described in Supplementary Text S3. The counterfactual approach is based on conceptualising ‘potential outcomes’ that would have been observed if certain conditions were met, regardless of the conditions that were in fact met (VanderWeele, 2016).

This method also allowed us to account for both intermediate confounding (i.e., confounding of the mediator-outcome relationship that may itself be affected by the exposure) by earlier offspring substance use and include a parental smoking-offspring depression interaction.

The Monte Carlo (MC) sample size was set to 100,000 to minimise fluctuations, and 95% confidence intervals were estimated using standard errors from 100 non-parametric bootstrap resamples. In large samples like ALSPAC, reducing bootstrap samples normally has little impact on parameter estimates and can reduce computational burden, while stability is heavily influenced by the MC sample size.

Mediation effects are presented as marginal odds ratios. Models both unadjusted and adjusted for the listed confounders were fitted. We also fitted models with and without adjusting for offspring substance use at age 13 as an intermediate confounder. We hypothesised that depressive symptoms may modify the effect of parental smoking on later offspring smoking and therefore included an exposure-mediator interaction in all mediation models. When such an interaction is present, the direct and indirect effects are no longer constant across levels of parental smoking. Consequently, the total effect is decomposed into the pure natural direct effect (PNDE) and total natural indirect effect (TNIE) as these effects remain well defined in the presence of effect modification by the mediator (Supplementary Text S3). In the g-computation mediation models, interactions are incorporated on the risk scale.

### Regression analysis

Standard unadjusted and adjusted logistic regression models were used to investigate the observed associations along each pathway (shown in Figure 1) providing an empirical summary of the relationships between variables. We also fit models including a parental smoking-offspring depressive symptoms interaction that is estimated multiplicatively on the odds ratio scale.

## Results

### Participant characteristics

There was a total of 6,729 participants in the analytic sample and 1,050 (16%) had complete data on all variables included in this analysis.

In the analytic sample 27% of participants had at least one parent who smoked daily at age 12. In those with available data, 22% reported higher depressive symptoms at age 14, 19% reported currently smoking at age 16, and 16% had used substances in past 6 months at age 13.

Most confounders showed some differences between those exposed and unexposed to parental smoking. Sex differences were observed between participants with lower and higher depressive symptoms in the analytic sample. Other sociodemographic factors such as ethnicity, parental social class and education, neighbourhood quality and deprivation, household income, and housing tenure showed limited variation by offspring depression (Table 1).

**Table 1.** Descriptive statistics in those with available data in the analytic sample (n = 6,729)

| Characteristic | Overall |  | Parental smoking (12y) |  | Offspring depression (14y) |  |
| --- | --- | --- | --- | --- | --- | --- |
|  | Missing | N = 6,729<sup>a</sup> | No N = 4,908<sup>a</sup> | Yes N = 1,821<sup>a</sup> | <8 N = 3,702<sup>a</sup> | ≥8 N = 1,034<sup>a</sup> |
| **Parental daily smoking (12y)** | 0 (0%) |  |  |  |  |  |
| No |  | 4,908 (73%) |  |  | 2,852 (77%) | 746 (72%) |
| Yes |  | 1,821 (27%) |  |  | 850 (23%) | 288 (28%) |
| **Offspring substance use (13y)** | 1,489 (22%) |  |  |  |  |  |
| No |  | 4,384 (84%) | 3,347 (86%) | 1,037 (78%) | 3,061 (86%) | 747 (76%) |
| Yes |  | 856 (16%) | 555 (14%) | 301 (22%) | 479 (14%) | 230 (24%) |
| **Offspring SMFQ<sup>b</sup> scores (14y)** | 1,993 (30%) |  |  |  |  |  |
| <8 |  | 3,702 (78%) | 2,852 (79%) | 850 (75%) |  |  |
| ≥8 |  | 1,034 (22%) | 746 (21%) | 288 (25%) |  |  |
| **Offspring current smoking (16y)** | 2,649 (39%) |  |  |  |  |  |
| No |  | 3,305 (81%) | 2,620 (84%) | 685 (72%) | 2,125 (84%) | 539 (74%) |
| Yes |  | 775 (19%) | 502 (16%) | 273 (28%) | 411 (16%) | 188 (26%) |
| **Mother smoked in pregnancy (0y)** | 472 (7.0%) |  |  |  |  |  |
| No |  | 5,019 (80%) | 4,221 (92%) | 798 (47%) | 2,911 (85%) | 748 (78%) |
| Yes |  | 1,238 (20%) | 345 (7.6%) | 893 (53%) | 525 (15%) | 209 (22%) |
| **Oldest parent's age at birth** | 2,172 (32%) | 31.6 (5.3) | 32.0 (5.1) | 30.6 (5.8) | 31.8 (5.3) | 32.2 (5.2) |
| **Lowest parental social class (0y)** | 679 (10%) |  |  |  |  |  |
| I/II |  | 1,851 (31%) | 1,517 (34%) | 334 (21%) | 1,080 (32%) | 286 (31%) |
| III |  | 3,229 (53%) | 2,360 (53%) | 869 (55%) | 1,817 (54%) | 486 (53%) |
| IV/V |  | 970 (16%) | 605 (13%) | 365 (23%) | 481 (14%) | 150 (16%) |
| **Lowest parental education (0y)** | 581 (8.6%) |  |  |  |  |  |
| A level or above |  | 2,119 (34%) | 1,777 (39%) | 342 (21%) | 1,257 (37%) | 346 (37%) |
| O level |  | 2,120 (34%) | 1,556 (34%) | 564 (35%) | 1,186 (35%) | 324 (34%) |
| Vocational/CSE |  | 1,909 (31%) | 1,224 (27%) | 685 (43%) | 971 (28%) | 273 (29%) |
| **Neighbourhood quality index (0y)** | 683 (10%) | 8.33 (2.13) | 8.48 (2.06) | 7.91 (2.24) | 8.42 (2.07) | 8.31 (2.13) |
| **Townsend deprivation score quintiles (8y)** | 1,230 (18%) |  |  |  |  |  |
| Q1 |  | 2,010 (37%) | 1,616 (40%) | 394 (28%) | 1,226 (39%) | 328 (36%) |
| Q2 |  | 949 (17%) | 707 (17%) | 242 (17%) | 541 (17%) | 168 (19%) |
| Q3 |  | 1,054 (19%) | 762 (19%) | 292 (21%) | 602 (19%) | 164 (18%) |
| Q4 |  | 1,069 (19%) | 752 (18%) | 317 (22%) | 605 (19%) | 175 (19%) |
| Q5 |  | 417 (7.6%) | 242 (5.9%) | 175 (12%) | 200 (6.3%) | 70 (7.7%) |
| **Household income per week (8y)** | 1,428 (21%) |  |  |  |  |  |
| 400+ |  | 2,849 (54%) | 2,328 (59%) | 521 (38%) | 1,725 (57%) | 463 (54%) |

|  | Overall |  | Parental smoking (12y) |  | Offspring depression (14y) |  |
| --- | --- | --- | --- | --- | --- | --- |
| Characteristic | Missing | N = 6,729<sup>a</sup> | No N = 4,908<sup>a</sup> | Yes N = 1,821<sup>a</sup> | <8 N = 3,702<sup>a</sup> | ≥8 N = 1,034<sup>a</sup> |
| <400 |  | 2,452 (46%) | 1,609 (41%) | 843 (62%) | 1,309 (43%) | 391 (46%) |
| **Housing tenure (10y)** | 542 (8.1%) |  |  |  |  |  |
| *Mortgaged/Owned* |  | 5,461 (88%) | 4,209 (92%) | 1,252 (77%) | 3,160 (90%) | 869 (89%) |
| *Rented/Other* |  | 726 (12%) | 361 (7.9%) | 365 (23%) | 337 (9.6%) | 108 (11%) |
| **Highest parental EPDS<sup>c</sup> score (8y)** | 3,345 (50%) | 7.2 (5.1) | 7.0 (4.9) | 7.7 (5.6) | 6.9 (4.9) | 7.9 (5.0) |
| **Parental mental health condition (8y)** | 2,902 (43%) |  |  |  |  |  |
| *No* |  | 2,778 (73%) | 2,186 (76%) | 592 (63%) | 1,672 (75%) | 458 (71%) |
| *Yes* |  | 1,049 (27%) | 705 (24%) | 344 (37%) | 546 (25%) | 186 (29%) |
| **Sex assigned at birth** | 0 (0%) |  |  |  |  |  |
| *Male* |  | 3,346 (50%) | 2,437 (50%) | 909 (50%) | 1,984 (54%) | 333 (32%) |
| *Female* |  | 3,383 (50%) | 2,471 (50%) | 912 (50%) | 1,718 (46%) | 701 (68%) |
| **Ethnicity** | 515 (7.7%) |  |  |  |  |  |
| *White* |  | 5,992 (96%) | 4,456 (97%) | 1,536 (95%) | 3,328 (96%) | 915 (96%) |
| *Ethnically minoritised* |  | 222 (3.6%) | 140 (3.0%) | 82 (5.1%) | 121 (3.5%) | 34 (3.6%) |
| **Physical/emotional abuse (9y)** | 800 (12%) |  |  |  |  |  |
| *No* |  | 5,739 (97%) | 4,290 (97%) | 1,449 (95%) | 3,273 (97%) | 909 (96%) |
| *Yes* |  | 190 (3.2%) | 116 (2.6%) | 74 (4.9%) | 92 (2.7%) | 40 (4.2%) |
| **Peer substance use (13y)** | 1,544 (23%) |  |  |  |  |  |
| *No* |  | 2,627 (51%) | 2,077 (54%) | 550 (42%) | 1,925 (55%) | 397 (41%) |
| *Yes* |  | 2,558 (49%) | 1,784 (46%) | 774 (58%) | 1,568 (45%) | 576 (59%) |
| **Parental monitoring (13y)** | 2,343 (35%) |  |  |  |  |  |
| *Always/most of the time* |  | 3,704 (84%) | 2,794 (86%) | 910 (81%) | 2,566 (88%) | 642 (78%) |
| *Never/Hardly ever/Sometimes* |  | 682 (16%) | 469 (14%) | 213 (19%) | 365 (12%) | 176 (22%) |
| **Experienced any bullying (13y)** | 1,438 (21%) |  |  |  |  |  |
| *No* |  | 2,687 (51%) | 2,055 (52%) | 632 (47%) | 1,982 (56%) | 325 (33%) |
| *Yes* |  | 2,604 (49%) | 1,881 (48%) | 723 (53%) | 1,581 (44%) | 665 (67%) |

<sup>a</sup> n (%) or Mean (SD) based on available data for each variable; <sup>b</sup> Short Mood and Feelings Questionnaire; <sup>c</sup> Edinburgh Postnatal Depression Scale

Similar differences were observed when looking at the full ALSPAC sample (Supplementary Table S1). There were also similar correlations between variables in the analytic (Supplementary Figure S2) and full ALSPAC sample (Supplementary Figure S3).

### Missing data

Many of the analysis variables, including the main outcome (offspring smoking at age 16), were associated with missingness (Supplementary Table S2) and being in the analytic sample (Supplementary Table S3) in univariable models. This reflects prior studies showing that loss to follow-up in ALSPAC is related to socioeconomic position (Howe, Tilling, Galobardes, & Lawlor, 2013) and polygenic scores for smoking (Taylor et al., 2018). However, after adjusting for parental smoking, offspring depression and a subset of key confounders offspring smoking was no longer associated with missingness (Supplementary Table S4). This indicates that complete-records analysis (CRA) is likely to be unbiased (Hughes, Heron, Sterne, & Tilling, 2019). Since missingness may be plausibly explained by observed variables and auxiliary data were available at earlier, more complete, waves of data collection, multiple imputation (MI) is likely to provide more efficient estimates than CRA in this setting.

Patterns of missingness in the analytic sample are shown (Supplementary Figure S4). Parental mental health and age measures rely on partner-based questionnaire data and so show greater missingness than other parental variables. The main outcome, current smoking at age 16, also shows a high proportion of missingness given this is the latest collected measure used in this analysis and attrition in ALSPAC has occurred over time (Boyd et al., 2013).

For the primary exposure regression coefficients, the Monte Carlo error was <10% of its standard error, indicating that 50 imputations were sufficient. For some confounders errors were slightly higher (<12%) but still reasonable.

Complete-records analyses showed limited evidence of effects (Supplementary Tables S5–S8) and associations were generally weaker and less consistent than in the MI analyses. This likely reflects reduced precision due to the smaller sample size.

### Mediation analysis

In unadjusted analyses there was evidence for a total causal effect (TCE) of parental smoking at age 12 on offspring current smoking at age 16 (OR = 2.16, 95% CI = 1.81-2.57). Most of the total effect encompassed the pure natural direct effect (PNDE; OR = 2.09, 95% CI = 1.75-2.49) but there was evidence for a small total natural indirect effect (TNIE) via depressive symptoms (OR = 1.03, 95% CI = 1.01-1.06), with a proportion mediated of around 4%. The controlled direct effect (CDE) shows the effect of parental smoking on offspring smoking if depressive symptoms were intervened on and held constant (at a low level) for everyone. Here the CDE (OR = 2.13, 95% CI = 1.73-2.62) was slightly higher than the PNDE, which instead holds depressive symptoms at the natural level they would take under no parental smoking.

In fully adjusted analyses, there was a smaller TCE of parental smoking on offspring smoking (OR = 1.44, 95% CI = 1.19-1.73). The PNDE (OR = 1.43, 95% CI = 1.19–1.73) was almost identical to the total effect and the TNIE via depressive symptoms was null (OR = 1.00, 95% CI = 0.99–1.01). This suggests there was little evidence that the intergenerational smoking effect is meaningfully mediated by offspring depressive symptoms.

The CDE (OR = 1.46, 95% CI = 1.18-1.81) gave a similar estimate. This suggests that the effect of the parental smoking on offspring smoking is not due to mediation by, or interaction with, offspring depressive symptoms.

Similar results were observed after further adjusting for potential intermediate confounding by earlier offspring substance use (Table 2).

**Table 2.** G-computation formula estimates of the total and direct effects of parental smoking on offspring smoking and indirect effects via offspring depressive symptoms (n = 6,729)

|  | Unadjusted for baseline & mediator-outcome confounders |  |  | Adjusted for baseline & mediator-outcome confounders |  |  |
| --- | --- | --- | --- | --- | --- | --- |
| Effect | OR | LCI | UCI | OR | LCI | UCI |
| **No intermediate confounder** |  |  |  |  |  |  |
| TCE | 2.16 | 1.81 | 2.57 | 1.44 | 1.19 | 1.73 |
| CDE | 2.13 | 1.73 | 2.62 | 1.46 | 1.18 | 1.81 |
| PNDE | 2.09 | 1.75 | 2.49 | 1.43 | 1.19 | 1.73 |
| TNIE | 1.03 | 1.01 | 1.06 | 1.00 | 0.99 | 1.01 |
| **Including early offspring substance use (13y)** |  |  |  |  |  |  |
| TCE | 2.16 | 1.83 | 2.56 | 1.44 | 1.20 | 1.72 |
| CDE | 2.13 | 1.76 | 2.59 | 1.45 | 1.18 | 1.78 |
| PNDE | 2.10 | 1.77 | 2.49 | 1.43 | 1.19 | 1.72 |
| TNIE | 1.03 | 1.00 | 1.06 | 1.00 | 0.99 | 1.01 |
| Abbreviations: OR = Odds Ratio, LCI = Lower Confidence Interval, UCI = Upper Confidence Interval, TCE = Total causal effect, CDE = Controlled direct effect, PNDE = Pure Natural Direct Effect, TNIE = Total Natural Indirect Effect |  |  |  |  |  |  |

### Regression analysis

Unadjusted logistic regression analyses show that parental smoking at age 12 is associated with higher odds of both early offspring substance use (OR = 1.74, 95% CI = 1.49-2.02) at age 13 (Table 3), and higher offspring depressive symptoms (OR = 1.34, 95% CI = 1.14-1.57) at age 14 (Table 4). Early offspring substance use was also associated with later depressive symptoms (OR = 1.95, 95% CI = 1.65-2.31).

**Table 3.**
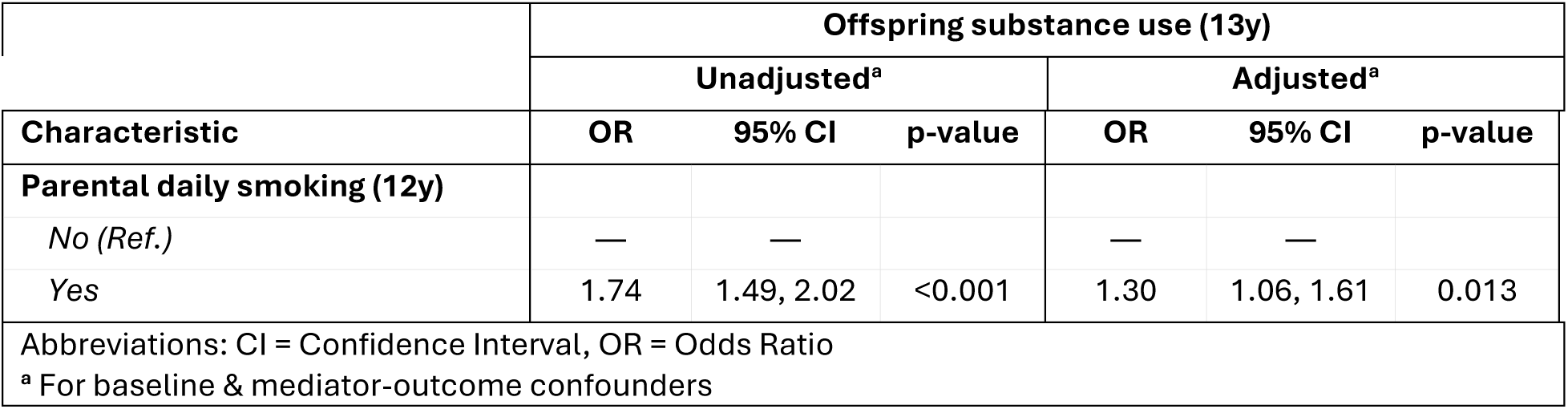
Logistic regression estimates showing associations of parental smoking on early offspring substance use (n = 6,729)

**Table 4.** Logistic regression estimates showing associations of parental smoking and early offspring substance use on offspring depressive symptoms (n = 6,729)

|  | Offspring depressive symptoms (14y) |  |  |  |  |  |
| --- | --- | --- | --- | --- | --- | --- |
|  | Unadjusted<sup>a</sup> |  |  | Adjusted<sup>a</sup> |  |  |
| Characteristic | OR | 95% CI | p-value | OR | 95% CI | p-value |
| **Parental daily smoking (12y)** |  |  |  |  |  |  |
| No (Ref.) | — | — |  | — | — |  |
| Yes | 1.34 | 1.14, 1.57 | <0.001 | 1.05 | 0.86, 1.29 | 0.642 |
| **Offspring substance use (13y)<sup>b</sup>** |  |  |  |  |  |  |
| No (Ref.) | — | — |  | — | — |  |
| Yes | 1.95 | 1.65, 2.31 | <0.001 | 1.30 | 1.06, 1.60 | 0.012 |
| Abbreviations: CI = Confidence Interval, OR = Odds Ratio |  |  |  |  |  |  |
| <sup>a</sup> For baseline & mediator-outcome confounders |  |  |  |  |  |  |
| <sup>b</sup> Adjusted for parental smoking at offspring at age 12 |  |  |  |  |  |  |

In fully adjusted analyses there was still evidence that parental smoking was associated with early offspring substance use (OR = 1.30, 95% CI = 1.06-1.61), and that offspring substance use was associated with higher depressive symptoms (OR = 1.30, 95% CI = 1.06-1.60), albeit with lower odds. However, the association between parental smoking and offspring depression was attenuated by further adjustment (OR = 1.05, 95% CI = 0.86-1.29).

Unadjusted regression analyses also showed that parental smoking at age 12 (OR = 2.16, 95% CI = 1.82-2.56), offspring substance use at age 13 (OR = 4.13, 95% CI = 3.37-5.07), and higher offspring depressive symptoms at age 14 (OR = 1.81, 95% CI = 1.45-2.25) all were associated with an increase in the odds of current offspring smoking at age 16 (Table 5).

**Table 5.** Logistic regression estimates showing associations of parental daily smoking, early offspring substance use and offspring depressive symptoms on later offspring smoking (n = 6,729)

|  | Offspring smoking (16y) |  |  |  |  |  |
| --- | --- | --- | --- | --- | --- | --- |
|  | Unadjusted<sup>a</sup> |  |  | Adjusted<sup>a</sup> |  |  |
| Characteristic | OR | 95% CI | p-value | OR | 95% CI | p-value |
| **Parental daily smoking (12y)** |  |  |  |  |  |  |
| No (Ref.) | — | — |  | — | — |  |
| Yes | 2.16 | 1.82, 2.56 | <0.001 | 1.49 | 1.21, 1.83 | <0.001 |
| **Offspring substance use (13y)<sup>b</sup>** |  |  |  |  |  |  |
| No (Ref.) | — | — |  | — | — |  |
| Yes | 4.13 | 3.37, 5.07 | <0.001 | 2.81 | 2.17, 3.62 | <0.001 |
| **Offspring SMFQ scores (14y)<sup>b</sup>** |  |  |  |  |  |  |
| <8 (Ref.) | — | — |  | — | — |  |
| ≥8 | 1.81 | 1.45, 2.25 | <0.001 | 1.35 | 1.06, 1.71 | 0.015 |
| **Offspring SMFQ scores (14y)<sup>b,c</sup>** |  |  |  |  |  |  |
| <8 (Ref.) | — | — |  | — | — |  |
| ≥8 | 1.59 | 1.26, 2.00 | <0.001 | 1.30 | 1.02, 1.66 | 0.036 |

Abbreviations: CI = Confidence Interval, OR = Odds Ratio
<sup>a</sup> For baseline & mediator-outcome confounders
<sup>b</sup> Adjusted for parental smoking at offspring at age 12
<sup>c</sup> Adjusted for offspring substance use at age 13

Further adjusting for earlier offspring substance use partially attenuated the association of offspring depressive symptoms with later current smoking (OR = 1.59, 95% CI = 1.26-2.00).

Fully adjusted analyses also show associations between parental smoking (OR = 1.49, 95% CI = 1.21-1.83), early substance use (OR = 2.81, 95% CI = 2.17-3.62), and offspring depression (OR = 1.35, 95% CI = 1.06-1.71) with later offspring smoking, with smaller effects. The association between offspring depression with later offspring smoking (OR = 1.30, 95% CI = 1.02-1.66) was only slightly attenuated in fully adjusted models when adjusting for offspring substance use.

There was no clear evidence for a parental smoking-offspring depression interaction on offspring smoking (Supplementary Table S9) in either unadjusted (OR = 0.96, 95% CI = 0.67-1.39) or adjusted (OR = 0.94, 95% CI = 0.65-1.38) analyses.

Together the regression results reflect the mediation analyses: there was a strong association between parental smoking and offspring smoking, but there was a lack of evidence to suggest parental smoking was associated with offspring depressive symptoms when adjusting for confounding.

## Discussion

We examined the intergenerational transmission of smoking behaviours, investigating whether offspring depression mediates the effect of parental daily smoking in early adolescence on offspring current smoking in late adolescence. Using data from a large longitudinal birth cohort, we show a strong direct effect of parental smoking on offspring smoking. However, we found little evidence that this effect was mediated via high offspring depressive symptoms after adjusting for confounding. The controlled direct effect was also the same as the total effect, which suggests that even if it were possible to eliminate high depression symptoms, this would have little impact on the effect of parental smoking on offspring smoking.

Our results support existing literature showing that early substance use and depressive symptoms both independently predict later smoking (Chaiton et al., 2009; Fluharty et al., 2017; Hockenberry et al., 2011), and confirm that parental smoking is an important causal risk factor for later smoking in offspring (Hockenberry et al., 2011; Alves et al., 2017; Vitória et al., 2020).

However, our findings do not suggest that the latter operates through offspring depression. This contrasts with previous research suggesting that aspects of adolescent mental health may moderate the intergenerational transmission of smoking. For example, one study found that higher mental health competence mitigated the increased risk of smoking initiation among teenagers with smoking parents (Pearce et al., 2021). However, this study focused on skills-based components of positive mental health rather than low mood, and solely examined moderation rather than mediation, which may explain the apparent discrepancy with our findings. Several other observational studies have reported that parental, particularly maternal prenatal, smoking is associated with an increased risk of adolescent depression (Menezes et al., 2013), sometimes showing dose-response patterns. Related research has linked second-hand smoke exposure with higher depressive symptoms in adolescents (Jacob et al., 2020; Kim, Park, Choi, Lee, & Kim, 2016), and parental depression has been shown to moderate associations between adolescents’ depressive symptoms and their own smoking or vaping (Cho, 2019), consistent with the idea that family mental health context shapes smoking uptake. A sibling-design study further estimated controlled direct effects after accounting for the pathway through the offspring’s own smoking, finding some evidence of a direct effect of maternal smoking during pregnancy on the risk of depression (Shenassa, Rogers, & Buka, 2023).

Conversely, studies employing cross-cohort negative-control and discordant-sibling designs have found that the associations between parental smoking and offspring depression attenuate substantially after accounting for shared familial factors (Taylor et al., 2017). Similar conclusions have been reached by other work showing that links between parental smoking and offspring psychiatric outcomes, including psychosis, depression and anxiety disorders, are highly susceptible to residual confounding (Sarala et al., 2022; Taylor et al., 2014; Thapar & Rutter, 2009; Woolf et al., 2024). These findings suggest that associations between parental smoking and offspring depression are more plausibly non-causal, reflecting shared social, familial, or genetic vulnerabilities rather than a direct developmental pathway. This is consistent with our results and help to explain why depression did not mediate the parental-offspring smoking relationship in our study.

### Strengths and limitations

The first strength of this study is the use of multiple imputation to improve power and reduce potential bias due to missing data. Second, we employed a counterfactual mediation framework while thoroughly accounting for a wide range of baseline confounders and intermediate confounding. Third, the longitudinal design allowed us to time order the analysis variables to refine our understanding of the causal mechanisms linking parental smoking and offspring smoking.

There are also some limitations. First, the observational design means that residual confounding and reverse causation cannot be completely ruled out, but we have tried to mitigate this with extensive adjustment and by using longitudinal data. Second, detecting interaction effects requires large sample sizes and high statistical power, which may have limited our ability to detect smaller effects. This is reflected in the wide confidence intervals for the interaction terms meaning moderation effects may have been missed.

Third, although multiple imputation was used to reduce bias due to missing data, this approach relies on largely untestable assumptions. However, we have reduced the risk of bias due to data missing not at random by using a large number of auxiliary variables that are predictive of the missing data in our imputation procedure. Fourth, the accuracy of self-reported smoking and mental health measures, particularly in adolescence, may be affected by social desirability bias and under-reporting. In addition, the timing of assessments may limit interpretation: depressive symptoms at age 14 may precede full onset for some individuals, and smoking at age 16 may not capture later uptake. However, adolescence is when initiation typically occurs, and the lower cut-off used for depressive symptoms increases sensitivity to early emotional difficulties. Fifth, parental smoking was measured at a single time point and may not fully represent exposure across childhood, potentially diluting observed associations. However, age 12 closely precedes the period when offspring begin experimenting with cigarettes, and prior work suggests that the influence of parental smoking on offspring increases during adolescence (Bricker, Peterson, Sarason, Andersen, & Rajan, 2007). Finally, parental mental health, among other potential intermediate confounders or mediators, was not measured at the specific time point needed to include in the mediation analysis, limiting our ability to fully disentangle the role of other factors.

### Implications

Overall, these findings strengthen the evidence for direct intergenerational transmission of smoking behaviours but do not support a mediating role for adolescent depressive symptoms. Future work could explore alternative mechanisms, such as modelling, peer influence, parental mental health, and normative beliefs around smoking, to better understand how parental smoking influences offspring behaviour. Interventions will also need to consider broader family and social contexts. Although previous evidence suggested that interventions could be streamlined to focus on mental health to prevent smoking among adolescents whose parents smoke, this evidence suggests that interventions should not focus solely on depression and should be multifaceted and focus also on other potential mediators.

## Conclusion

Our findings show a strong effect of parental smoking on offspring smoking but this relationship appears to act through mechanisms other than offspring depression.

## Data availability

The informed consent obtained from ALSPAC participants does not allow the data to be made available through any third party maintained public repository. Supporting data are available from ALSPAC on request under the approved proposal number, B3499 and B4347. Full instructions for applying for data access can be found here: http://www.bristol.ac.uk/alspac/researchers/access/. Please note that the study website contains details of all the data that is available through a fully searchable data dictionary and variable search tool: http://www.bristol.ac.uk/alspac/researchers/our-data/. Study data were collected and managed using REDCap electronic data capture tools hosted at the University of Bristol (Harris et al., 2009).

Data access for this project was granted (B3499/B4347) before this study. Datasets were created using syntax templates from the 14th of March 2025.

The proposed analysis of data was pre-registered on the Open Science Framework (OSF) here: https://osf.io/jq7gr

The code used for data analysis is available in a github repository here: https://github.com/alexandrayas/ALSPAC_CRUK_smkvap/tree/main/Parental%20smoking

## Ethical approval

Ethical approval for the study was obtained from the ALSPAC Law and Ethics Committee and Local Research Ethics Committees (https://www.bristol.ac.uk/alspac/researchers/research-ethics/). Informed consent for the use of all data collected was obtained from participants following the recommendations of the ALSPAC Ethics and Law Committee at the time. Participants can contact the study team at any time to retrospectively withdraw consent for their data to be used. Study participation is voluntary and during all data collection sweeps, information was provided on the intended use of data. The completion of a questionnaire, either on paper or online, was considered to be written consent from participants to use their data for research purposes. For the majority of tests undertaken during face to face visits, verbal consent was obtained from participants (both parents and children as appropriate) prior to the start of any data collection. However, some tests required the completion of a written consent form.

## Data Availability

The informed consent obtained from ALSPAC participants does not allow the data to be made available through any third party maintained public repository. Supporting data are available from ALSPAC on request under the approved proposal number, B3499 and B4347. Please note that the study website contains details of all the data that is available through a fully searchable data dictionary and variable search tool.

http://www.bristol.ac.uk/alspac/researchers/access/

https://www.bristol.ac.uk/alspac/researchers/our-data/

## Acknowledgements

We are extremely grateful to all the families who took part in this study, the midwives for their help in recruiting them, and the whole ALSPAC team, which includes data collection staff, data and administrations staff, technical managers and the technical staff with the Bristol Bioresource Laboratory, based within the University of Bristol.

## Author contributions

*Alexandria Andrayas*: Conceptualization (lead); formal analysis (lead); visualization (lead); writing—original draft preparation (lead); writing— review and editing (lead). *Elinor Curnow*: Conceptualization (equal); formal analysis (equal); supervision (equal); writing—review and editing (equal). *Jasmine Khouja*: Conceptualization (equal); formal analysis (equal); writing—review and editing (equal). *Jon Heron*: Conceptualization (equal); formal analysis (equal); supervision (equal); writing—review and editing (equal). *Hannah Jones*: Conceptualization (equal); formal analysis (equal); writing—review and editing (equal).

*Lindsey Hines*: Conceptualization (equal); writing—review and editing (equal). *Marcus Munafò*: Funding acquisition (equal); writing—review and editing (equal). *Gemma Hammerton*: Conceptualization (equal); formal analysis (lead); supervision (equal); writing—review and editing (equal). *Hannah Sallis*: Funding acquisition (lead); conceptualization (lead); supervision (equal); writing—review and editing (equal).

