## Supplementary Materials for "Can adolescent depressive symptoms act as a target for reducing intergenerational smoking transmission?"

*Supplementary Text S1. Confounding*

Information on included confounders

The term “ethnically minoritised” is used to refer to participants from all other ethnic groups combined when compared to participants with two white parents, using ‘minoritised’ in a UK context.

Parental social class was derived from the mother’s and her partner’s occupations using the 1991 UK Office of Population Censuses and Surveys classification based on Registrar General’s Social Class categorisation, now replaced by the National Statistics Socio-economic classification (Office for National Statistics, 2010).

In the UK, A-levels are academic qualifications usually taken between ages 16 and 18. O-levels were academic exams taken at ages 14-16 before leaving school, while vocational qualifications and the Certificate of Secondary Education (CSE) offered routes for students not pursuing advanced academic study. O-levels and CSEs were replaced by General Certificates of Secondary Education (GCSEs).

The neighbourhood quality index is a summation using questions related to the mother’s assessment of the neighbourhood as lively, friendly, noisy, clean, attractive, polluted or dirty, where a higher score corresponds to a higher quality neighbourhood.

Townsend deprivation score quintiles divide areas into five groups based on their level of deprivation calculated using four census variables including unemployment, car ownership, home ownership, and household overcrowding (Townsend, Phillimore, & Beattie, 1988) and was derived here from postcodes.

The Edinburgh Postnatal Depression Scale is a questionnaire originally used to screen for postnatal depression (Cox, Holden, & Sagovsky, 1987). Here a higher score helps identify parents experiencing more symptoms of depression. Parental mental health conditions refer to if at least one parent reported ever experiencing drug addiction, alcoholism, schizophrenia, anorexia nervosa, severe depression, or other psychiatric problems. Physical or emotional abuse refers to whether the mother reported they, or their partner, had been physically or emotionally cruel to her children in the 3 years prior to the questionnaire.

Peer substance use refers to whether the offspring participant’s friends had ever smoked, drank alcohol, or used cannabis.

Parental monitoring refers to the teenager’s reported frequency that carers know what they do in their free time.

Bullying refers to whether the teenager reported that someone had done at least one of the following to them; taken personal belongings without asking first, threatened or blackmailed, hit or beaten up, tricked, called nasty names, or done other bad things.

Confounding in mediation analyses

In mediation analyses, four assumptions are made with respect to confounding. These include no unmeasured confounders for any of the paths and no confounders for the association between the mediator and outcome which lie on the causal pathway from the exposure. In the parametric g-computation formula, exposure, mediator, and outcome models are specified with the same covariate set. This ensures consistency when simulating counterfactual outcomes under different intervention scenarios. This means here we assume that the same set of covariates confound all paths. In the presence of intermediate confounding, the assumption of no unaccounted mediator-outcome confounding is adapted to include conditioning on the intermediate confounder (De Stavola et al., 2015).

Confounding in regression analyses

Caution is required when directly comparing across logistic regression models, each with different sets of confounders, given the non-collapsibility of the odds ratio with a common binary outcome. Here adjusted regressions include both pre-exposure baseline confounders and mediator–outcome confounders (measured after exposure but prior to the mediator) in all models for consistency across analytic approaches. However, the intermediate confounder (early offspring substance use) is only adjusted for in mediator-outcome regression models.

*Supplementary Figure S1. Predictor matrix showing the variables included in the imputation model, and methods used to impute each, where data was imputed up to the sample of participants with data on parental smoking at age 12*

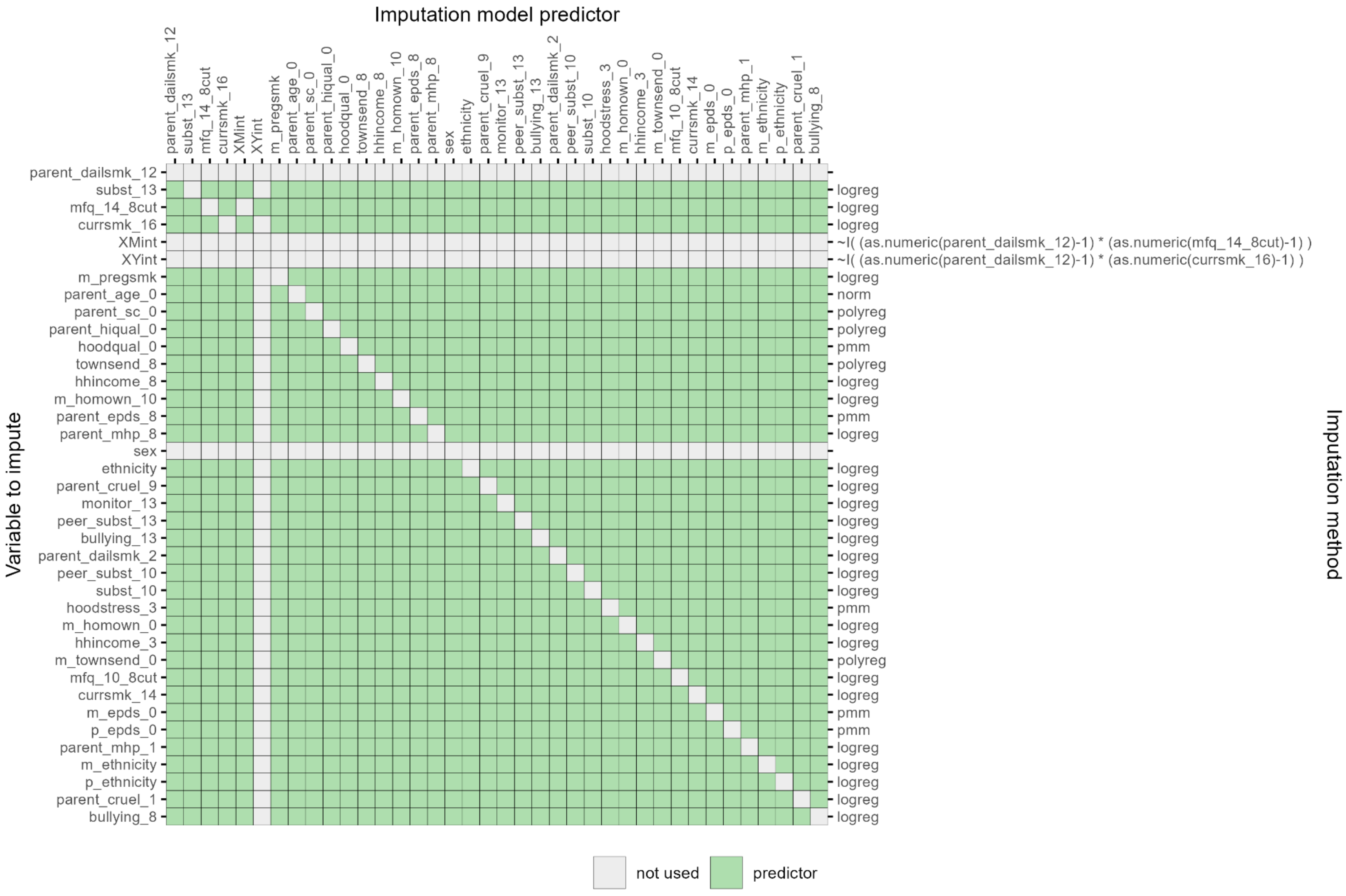

*Labels:* ***parent_dailsmk_12*** *= Parental daily smoking (12y);* ***subst_13*** *= Offspring substance use in past 6 months (13y);* ***mfq_14_8cut*** *= Offspring SMFQ scores; ≥8 (14y);* ***currsmk_16*** *= Current smoking (16y);* ***XMint*** *= Parental smoking-Offspring depression interaction term;* ***XYint*** *= Parental smoking-Offspring smoking interaction term;* ***m_pregsmk*** *= Mother smoked in pregnancy (0y);* ***parent_age_0*** *= Oldest parental age at birth;* ***parent_sc_0*** *= Lowest parental social class (0y);* ***parent_hiqual_0*** *= Lowest parental education (0y);* ***hoodqual_0*** *= Neighbourhood quality index (0y);* ***townsend_8*** *= Townsend deprivation score quintiles (8y);* ***hhincome_8*** *= Household income per week (8y);* ***m_homown_10*** *= Housing tenure (10y);* ***parent_epds_8*** *= Highest parental EPDS score (8y);* ***parent_mhp_8*** *= Parental mental health condition (8y);* ***sex*** *= Sex assigned at birth;* ***ethnicity*** *= Ethnicity;* ***parent_cruel_9*** *= Physical/emotional abuse (9y);* ***monitor_13*** *= Parental monitoring (13y);* ***peer_subst_13*** *= Peer substance use (13y);* ***bullying_13*** *= Experienced any bullying (13y);* ***parent_dailsmk_2*** *= Parental daily smoking (2y);* ***peer_subst_10*** *= Peer substance use (10y);* ***subst_10*** *= Offspring substance use in past 6 months (10y);* ***hoodstress_3*** *= Neighbourhood stress score (3y);* ***m_homown_0*** *= Housing tenure; rented/HA/Other (0y);* ***hhincome_3*** *= Household income per week; <200 (3y);* ***m_townsend_0*** *= Townsend deprivation score; quintiles (0y);* ***mfq_10_8cut*** *= Offspring SMFQ scores; ≥8 (10y);* ***currsmk_14*** *= Offspring current smoking (14y);* ***m_epds_0*** *= Maternal EPDS score (0y);* ***p_epds_0*** *= Paternal EPDS score (0y);* ***m_ethnicity*** *= Maternal ethnicity;* ***p_ethnicity*** *= Paternal ethnicity;* ***parent_mhp_1*** *= Parental mental health condition (1y);* ***parent_cruel_1*** *= Physical/emotional abuse (1y);* ***bullying_8*** *= Experienced any bullying (8y).*

*Supplementary Text S2. Imputation strategy and auxiliary variables*

We accounted for the exposure-mediator interaction as follows: since the outcome, exposure, and mediator here are all binary, and there is no missingness in the exposure, we include an exposure-mediator interaction for the outcome, and an exposure-outcome interaction for the mediator. The exposure-mediator interaction is also used to predict all other variables in the imputation model.

The auxiliary variables included in the imputation model are as follows: parental daily smoking at age 2, early offspring substance use, peer substance use and offspring SMFQ scores at age 10, offspring current smoking status at age 14, whether either parent had a mental health condition, and mother-reported emotional or physical abuse at age 1, average household income and neighbourhood stress score at age 3, bullying at age 8, and the following measures collected during pregnancy: parental ethnicity, maternal and parental EPDS scores, maternal home ownership, and Townsend deprivation score quintiles. These were included to increase the predictive ability of the imputation model and reduce potential bias due to data being missing not at random (MNAR).

*Supplementary Text S3. G-computation formula effects*

When there is not an exposure (X)-mediator (M) interaction, the total effect of X on the outcome (Y) can be decomposed into a natural direct effect (NDE), the effect of X on Y when the mediator M is fixed at its level under X=0, and a natural indirect effect (NIE), the portion of the effect operating through changes in M while X is held constant. Here the direct effect of X on Y is constant across levels of M. In this case, this could easily tell us how much of the effect of parental smoking on offspring smoking operates through depression.

However, when an X-M interaction is present, the direct effect of X on Y varies across levels of M, and the indirect effect may also depend on X. Therefore, the total effect cannot be decomposed into a single "direct" and "indirect" effect as, in our case, the indirect (mediated) pathway may be different for children of smokers vs non-smokers, and the direct effect of parental smoking depends on the child's depressive symptoms. Mediation is therefore heterogeneous and including an interaction violates the additivity required for the simple decomposition.

To overcome this, we instead report the pure natural direct effect (PNDE) and total natural indirect effect (TNIE). The PNDE represents the effect of changing X from 0 to 1 while fixing M at its value under X=0, isolating the direct pathway before M could change. The TNIE represents the effect of changing M from M(0) to M(1) while holding X=1, capturing the full indirect (M-mediated) pathway through M when X=1. In other words, the PNDE shows the effect of changing parental smoking while holding offspring depressive symptoms fixed at the value they would take if parents did not smoke. The TNIE shows the effect of changing depressive symptoms (as induced by changing parental smoking), while keeping parental smoking fixed as if all parents smoked.

When an interaction is present instead of interpreting mediation as "the proportion of the effect via depression", it instead shows how depressive symptoms contribute to the causal effect of parental on offspring smoking, under a decomposition that allows the mediator's effect to depend on the exposure status. In other words, how the offspring smoking risk might change if parental smoking changed depressive symptoms (TNIE), versus if depressive symptoms were held at their unexposed level (PNDE).

These effects remain well defined in the presence of effect modification by the mediator, but this cannot tell us how much the interaction contributes specifically to the direct and indirect effects.

**Total Causal Effect** (TCE): The value the outcome (*Y*),offspring smoking, would take if everybody had been exposed to parental smoking ($X$=1) versus everyone having no parents who smoke ($X$=0) where offspring depression varies naturally: $E[Y\{X=1,M(X=1)\}] - E[Y\{X=0,M(X=0)\}]$.

**Pure Natural Direct Effect** (PNDE): The direct (unmediated) effect of parental smoking on offspring smoking when offspring depressive symptoms takes the value it would take in the absence of parental smoking. In other words, the difference between potential outcomes if all had a parent who smokes but offspring depression is fixed at the level under no parental smoking, versus if everyone had no parents who smoke: $E[Y\{X=1, M(X=0)\}] - E[Y\{X=0, M(X=0)\}]$.

**Total Natural Indirect Effect** (TNIE): Difference between TCE and PNDE, capturing the effect of parental smoking on offspring smoking that operates by changing offspring depressive symptoms while keeping parental smoking fixed for everyone: $E[Y\{X=1, M(X=1)\}] - E[Y\{X=1, M(X=0)\}]$.

**Proportion mediated** (PM): Proportion of the total effect of parental smoking on offspring smoking mediated by high offspring depressive symptoms (TNIE / TCE).

**Controlled Direct Effect** (CDE): Difference between potential outcomes for parental smoking vs no parental smoking, if offspring depressive symptoms is intervened on and held constant, in this case fixed to low (m=0), for everyone: $CDE(m) = E[Y(X=1,M=m)] - E[Y(X=0,M=m)]$.

*Supplementary Table S1. Descriptive statistics in full ALSPAC sample with available data on sex*

| | Overall | | Parental smoking (12y) | | Offspring depression (14y) | |
|---|---|---|---|---|---|---|
| **Characteristic** | **Missing** | **N = 15,029**[a] | **No** N = 4,908[a] | **Yes** N = 1,821[a] | **<8** N = 4,672[a] | **≥8** N = 1,339[a] |
| **Parental daily smoking (12y)** | 8,300 (55%) | | | | | |
| *No* | | 4,908 (73%) | | | 2,852 (77%) | 746 (72%) |
| *Yes* | | 1,821 (27%) | | | 850 (23%) | 288 (28%) |
| **Offspring substance use (13y)** | 8,342 (56%) | | | | | |
| *No* | | 5,520 (83%) | 3,347 (86%) | 1,037 (78%) | 3,721 (86%) | 920 (75%) |
| *Yes* | | 1,167 (17%) | 555 (14%) | 301 (22%) | 622 (14%) | 299 (25%) |
| **Offspring SMFQ[b] scores (14y)** | 9,018 (60%) | | | | | |
| *<8* | | 4,672 (78%) | 2,852 (79%) | 850 (75%) | | |
| *≥8* | | 1,339 (22%) | 746 (21%) | 288 (25%) | | |
| **Offspring current smoking (16y)** | 9,963 (66%) | | | | | |
| *No* | | 4,061 (80%) | 2,620 (84%) | 685 (72%) | 2,457 (83%) | 628 (74%) |
| *Yes* | | 1,005 (20%) | 502 (16%) | 273 (28%) | 492 (17%) | 224 (26%) |
| **Mother smoked in pregnancy (0y)** | 2,729 (18%) | | | | | |
| *No* | | 8,661 (70%) | 4,221 (92%) | 798 (47%) | 3,529 (83%) | 933 (76%) |
| *Yes* | | 3,639 (30%) | 345 (7.6%) | 893 (53%) | 701 (17%) | 291 (24%) |
| **Oldest parent's age at birth** | 6,709 (45%) | 30.9 (5.7) | 32.0 (5.1) | 30.6 (5.8) | 31.7 (5.4) | 32.0 (5.3) |
| **Lowest parental social class (0y)** | 3,515 (23%) | | | | | |
| *I/II* | | 2,993 (26%) | 1,517 (34%) | 334 (21%) | 1,260 (30%) | 352 (30%) |
| *III* | | 6,212 (54%) | 2,360 (53%) | 869 (55%) | 2,242 (54%) | 605 (52%) |
| *IV/V* | | 2,309 (20%) | 605 (13%) | 365 (23%) | 632 (15%) | 212 (18%) |
| **Lowest parental education (0y)** | 3,261 (22%) | | | | | |
| *A level or above* | | 3,266 (28%) | 1,777 (39%) | 342 (21%) | 1,446 (35%) | 401 (33%) |
| *O level* | | 3,899 (33%) | 1,556 (34%) | 564 (35%) | 1,448 (35%) | 413 (34%) |
| *Vocational/CSE* | | 4,603 (39%) | 1,224 (27%) | 685 (43%) | 1,289 (31%) | 384 (32%) |
| **Neighbourhood quality index (0y)** | 2,346 (16%) | 8.09 (2.28) | 8.48 (2.06) | 7.91 (2.24) | 8.37 (2.10) | 8.20 (2.24) |
| **Townsend deprivation score quintiles (8y)** | 7,510 (50%) | | | | | |
| *Q1* | | 2,634 (35%) | 1,616 (40%) | 394 (28%) | 1,403 (38%) | 388 (36%) |
| *Q2* | | 1,268 (17%) | 707 (17%) | 242 (17%) | 629 (17%) | 191 (18%) |
| *Q3* | | 1,422 (19%) | 762 (19%) | 292 (21%) | 699 (19%) | 193 (18%) |
| *Q4* | | 1,542 (21%) | 752 (18%) | 317 (22%) | 748 (20%) | 213 (20%) |
| *Q5* | | 653 (8.7%) | 242 (5.9%) | 175 (12%) | 258 (6.9%) | 89 (8.3%) |
| **Household income per week (8y)** | 7,949 (53%) | | | | | |
| *400+* | | 3,602 (51%) | 2,328 (59%) | 521 (38%) | 1,935 (55%) | 531 (53%) |

| | Overall | | Parental smoking (12y) | | Offspring depression (14y) | |
|---|---|---|---|---|---|---|
| **Characteristic** | **Missing** | **N = 15,029**[a] | **No** N = 4,908[a] | **Yes** N = 1,821[a] | **<8** N = 4,672[a] | **≥8** N = 1,339[a] |
| *<400* | | 3,478 (49%) | 1,609 (41%) | 843 (62%) | 1,557 (45%) | 466 (47%) |
| **Housing tenure (10y)** | 6,931 (46%) | | | | | |
| *Mortgaged/Owned* | | 6,987 (86%) | 4,209 (92%) | 1,252 (77%) | 3,641 (90%) | 1,016 (88%) |
| *Rented/Other* | | 1,111 (14%) | 361 (7.9%) | 365 (23%) | 423 (10%) | 133 (12%) |
| **Highest parental EPDS[c] score (8y)** | 10,764 (72%) | 7.4 (5.2) | 7.0 (4.9) | 7.7 (5.6) | 7.1 (5.0) | 8.0 (5.2) |
| **Parental mental health condition (8y)** | 10,059 (67%) | | | | | |
| *No* | | 3,478 (70%) | 2,186 (76%) | 592 (63%) | 1,878 (75%) | 518 (69%) |
| *Yes* | | 1,492 (30%) | 705 (24%) | 344 (37%) | 642 (25%) | 234 (31%) |
| **Sex assigned at birth** | 0 (0%) | | | | | |
| *Male* | | 7,683 (51%) | 2,437 (50%) | 909 (50%) | 2,504 (54%) | 432 (32%) |
| *Female* | | 7,346 (49%) | 2,471 (50%) | 912 (50%) | 2,168 (46%) | 907 (68%) |
| **Ethnicity** | 2,940 (20%) | | | | | |
| *White* | | 11,479 (95%) | 4,456 (97%) | 1,536 (95%) | 4,073 (96%) | 1,167 (96%) |
| *Ethnically minoritised* | | 610 (5.0%) | 140 (3.0%) | 82 (5.1%) | 166 (3.9%) | 45 (3.7%) |
| **Physical/emotional abuse (9y)** | 7,172 (48%) | | | | | |
| *No* | | 7,592 (97%) | 4,290 (97%) | 1,449 (95%) | 3,791 (97%) | 1,077 (96%) |
| *Yes* | | 265 (3.4%) | 116 (2.6%) | 74 (4.9%) | 111 (2.8%) | 45 (4.0%) |
| **Peer substance use (13y)** | 8,392 (56%) | | | | | |
| *No* | | 3,249 (49%) | 2,077 (54%) | 550 (42%) | 2,305 (54%) | 487 (40%) |
| *Yes* | | 3,388 (51%) | 1,784 (46%) | 774 (58%) | 1,987 (46%) | 731 (60%) |
| **Parental monitoring (13y)** | 9,359 (62%) | | | | | |
| *Always/most of the time* | | 4,749 (84%) | 2,794 (86%) | 910 (81%) | 3,173 (87%) | 787 (77%) |
| *Never/Hardly ever/Sometimes* | | 921 (16%) | 469 (14%) | 213 (19%) | 477 (13%) | 236 (23%) |
| **Experienced any bullying (13y)** | 8,265 (55%) | | | | | |
| *No* | | 3,380 (50%) | 2,055 (52%) | 632 (47%) | 2,404 (55%) | 398 (32%) |
| *Yes* | | 3,384 (50%) | 1,881 (48%) | 723 (53%) | 1,971 (45%) | 838 (68%) |

[a] n (%) or Mean (SD) based on available data for each variable; [b] Short Mood and Feelings Questionnaire; [c] Edinburgh Postnatal Depression Scale

*Supplementary Figure S2. Correlation matrix in analytic sample with data on parental smoking (n = 6,741)*

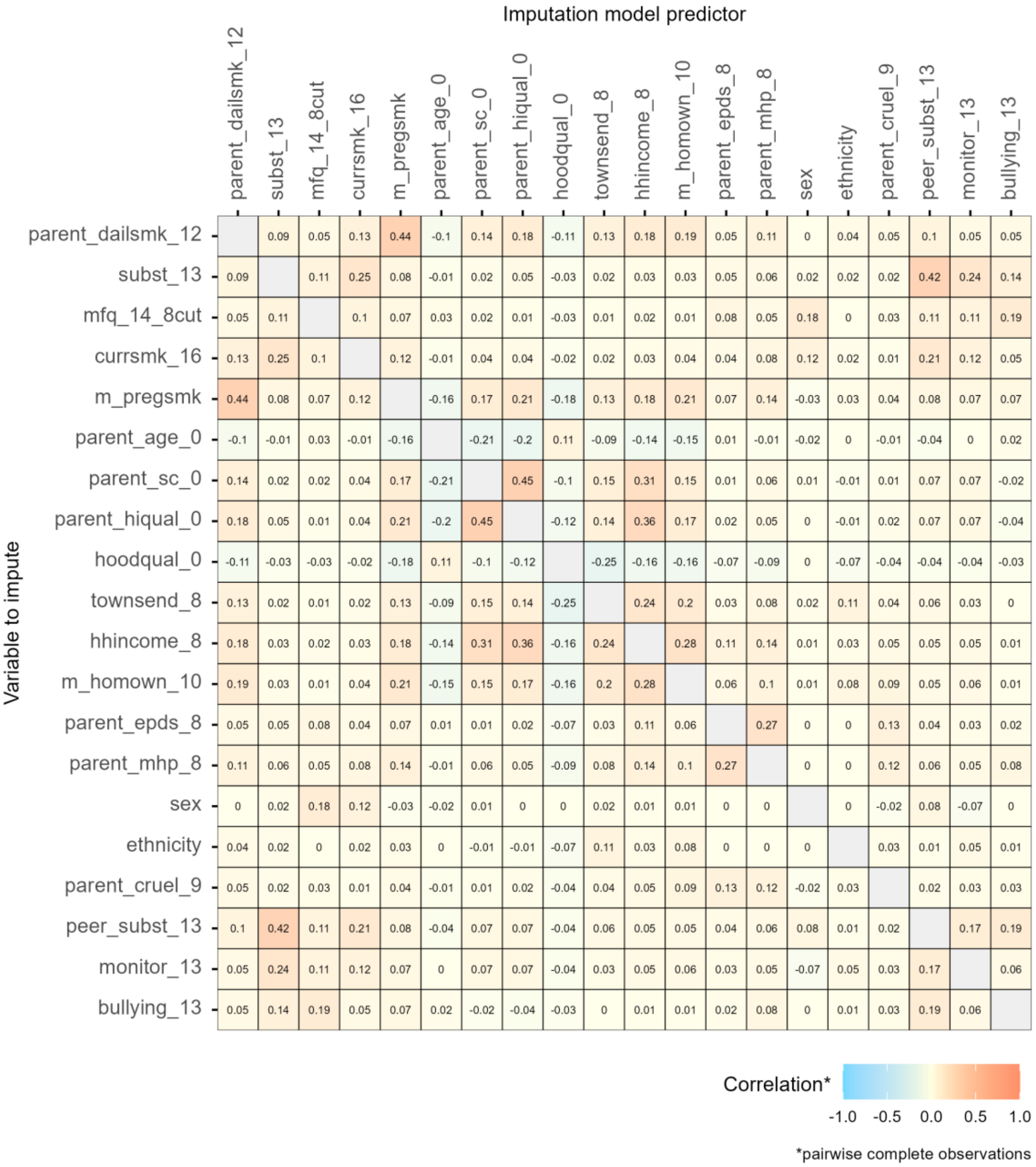


*Labels: **parent_dailsmk_12** = Parental daily smoking (12y); **subst_13** = Offspring substance use in past 6 months (13y); **mfq_14_8cut** = Offspring SMFQ scores; ≥8 (14y); **currsmk_16** = Offspring current smoking (16y); **XMint** = Parental smoking-Offspring depression interaction term; **XYint** = Parental smoking-Offspring smoking interaction term; **m_pregsmk** = Mother smoked in pregnancy (0y); **parent_age_0** = Oldest parental age at birth; **parent_sc_0** = Lowest parental social class (0y); **parent_hiqual_0** = Lowest parental education (0y); **hoodqual_0** = Neighbourhood quality index (0y); **townsend_8** = Townsend deprivation score quintiles (8y); **hhincome_8** = Household income per week (8y); **m_homown_10** = Housing tenure (10y); **parent_epds_8** = Highest parental EPDS score (8y); **parent_mhp_8** = Parental mental health condition (8y); **sex** = Sex assigned at birth; **ethnicity** = Ethnicity; **parent_cruel_9** = Physical/emotional abuse (9y); **monitor_13** = Parental monitoring (13y); **peer_subst_13** = Peer substance use (13y); **bullying_13** = Experienced any bullying (13y).*

*Supplementary Figure S3. Correlation matrix in full ALSPAC sample (n = 15,029)*

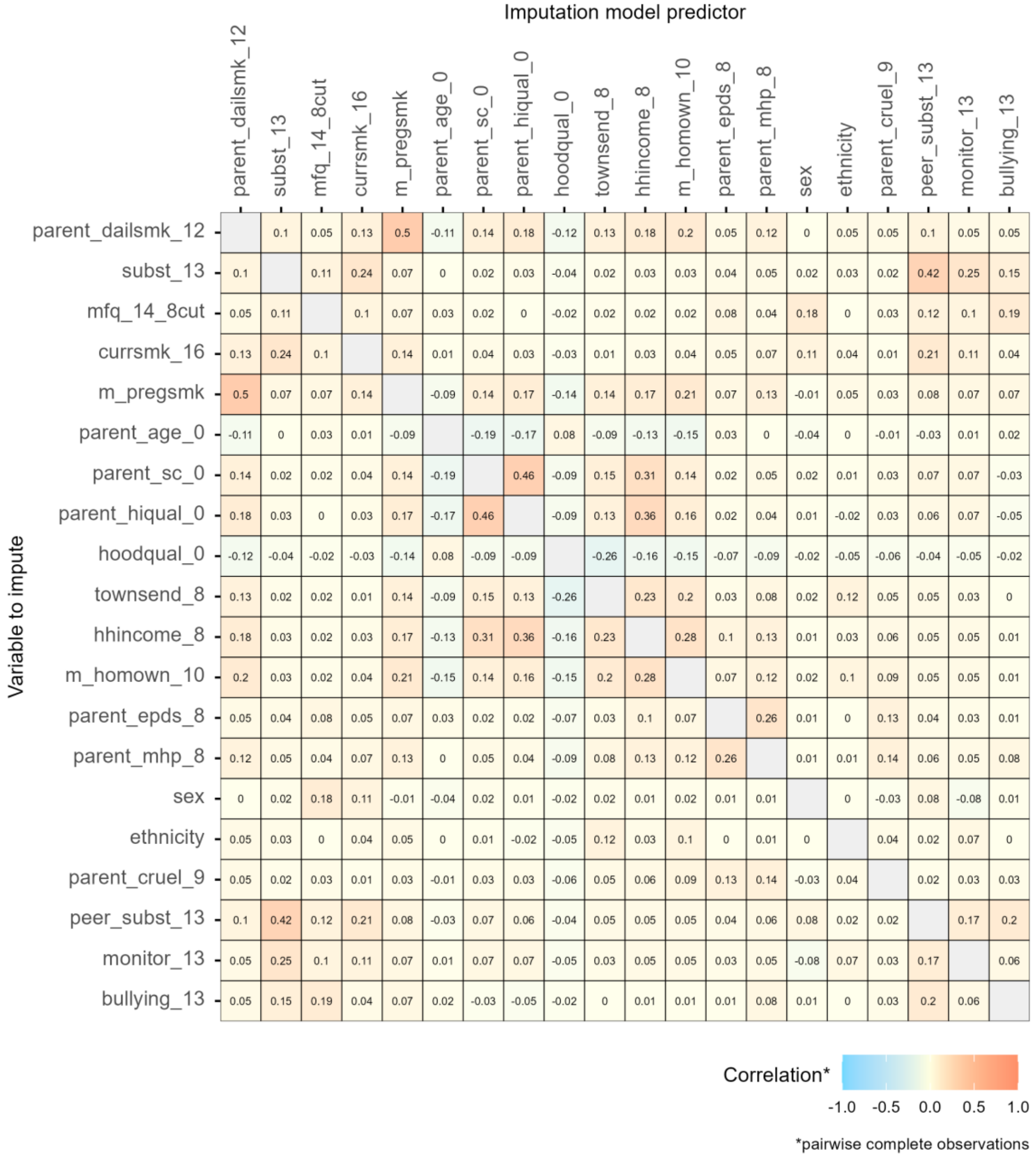


*Labels: **parent_dailsmk_12** = Parental daily smoking (12y); **subst_13** = Offspring substance use in past 6 months (13y); **mfq_14_8cut** = Offspring SMFQ scores; ≥8 (14y); **currsmk_16** = Offspring current smoking (16y); **XMint** = Parental smoking-Offspring depression interaction term; **XYint** = Parental smoking-Offspring smoking interaction term; **m_pregsmk** = Mother smoked in pregnancy (0y); **parent_age_0** = Oldest parental age at birth; **parent_sc_0** = Lowest parental social class (0y); **parent_hiqual_0** = Lowest parental education (0y); **hoodqual_0** = Neighbourhood quality index (0y); **townsend_8** = Townsend deprivation score quintiles (8y); **hhincome_8** = Household income per week (8y); **m_homown_10** = Housing tenure (10y); **parent_epds_8** = Highest parental EPDS score (8y); **parent_mhp_8** = Parental mental health condition (8y); **sex** = Sex assigned at birth; **ethnicity** = Ethnicity; **parent_cruel_9** = Physical/emotional abuse (9y); **monitor_13** = Parental monitoring (13y); **peer_subst_13** = Peer substance use (13y); **bullying_13** = Experienced any bullying (13y).*

*Supplementary Table S2. Univariable associations between analysis variables and missingness (i.e. whether all data are reported or not, coded as 1=complete case, 0=missing from complete case sample) using the analytic sample*

| Characteristic | N | OR | 95% CI | p-value |
|---|---|---|---|---|
| **Parental daily smoking (12y)** | 6,729 | | | |
| *No (Ref.)* | | — | — | |
| *Yes* | | 0.55 | 0.47, 0.65 | <0.001 |
| **Offspring substance use (13y)** | 5,240 | | | |
| *No (Ref.)* | | — | — | |
| *Yes* | | 0.57 | 0.46, 0.70 | <0.001 |
| **Offspring SMFQ scores (14y)** | 4,736 | | | |
| *<8 (Ref.)* | | — | — | |
| *≥8* | | 0.88 | 0.74, 1.04 | 0.144 |
| **Offspring current smoking (16y)** | 4,080 | | | |
| *No (Ref.)* | | — | — | |
| *Yes* | | 0.73 | 0.61, 0.88 | 0.001 |
| **Mother smoked in pregnancy (0y)** | 6,257 | | | |
| *No (Ref.)* | | — | — | |
| *Yes* | | 0.46 | 0.38, 0.56 | <0.001 |
| **Oldest parent's age at birth** | 4,557 | 1.04 | 1.03, 1.05 | <0.001 |
| **Lowest parental social class (0y)** | 6,050 | | | |
| *I/II (Ref.)* | | — | — | |
| *III* | | 0.65 | 0.56, 0.75 | <0.001 |
| *IV/V* | | 0.38 | 0.30, 0.48 | <0.001 |
| **Lowest parental education (0y)** | 6,148 | | | |
| *A level or above (Ref.)* | | — | — | |
| *O level* | | 0.61 | 0.52, 0.71 | <0.001 |
| *Vocational/CSE* | | 0.38 | 0.31, 0.45 | <0.001 |
| **Neighbourhood quality index (0y)** | 6,046 | 1.07 | 1.04, 1.11 | <0.001 |
| **Townsend deprivation score quintiles (8y)** | 5,499 | | | |
| *Q1 (Ref.)* | | — | — | |
| *Q2* | | 0.79 | 0.65, 0.95 | 0.015 |
| *Q3* | | 0.71 | 0.58, 0.85 | <0.001 |
| *Q4* | | 0.71 | 0.58, 0.86 | <0.001 |
| *Q5* | | 0.47 | 0.34, 0.64 | <0.001 |
| **Household income per week (8y)** | 5,301 | | | |
| *400+ (Ref.)* | | — | — | |
| *<400* | | 0.49 | 0.42, 0.56 | <0.001 |
| Housing tenure (10y) | 6,187 | | | |
| *Mortgaged/Owned (Ref.)* | | — | — | |
| *Rented/Other* | | 0.41 | 0.31, 0.53 | <0.001 |

| Characteristic | N | OR | 95% CI | p-value |
|---|---|---|---|---|
| **Highest parental EPDS score (8y)** | 3,384 | 0.98 | 0.97, 1.00 | 0.012 |
| **Parental mental health condition (8y)** | 3,827 | | | |
| *No (Ref.)* | | — | — | |
| *Yes* | | 0.51 | 0.43, 0.61 | <0.001 |
| **Sex assigned at birth** | 6,729 | | | |
| *Male (Ref.)* | | — | — | |
| *Female* | | 1.24 | 1.08, 1.41 | 0.002 |
| **Ethnicity** | 6,214 | | | |
| *White (Ref.)* | | — | — | |
| *Ethnically minoritised* | | 0.61 | 0.39, 0.92 | 0.024 |
| **Physical/emotional abuse (9y)** | 5,929 | | | |
| *No (Ref.)* | | — | — | |
| *Yes* | | 0.70 | 0.44, 1.05 | 0.097 |
| **Peer substance use (13y)** | 5,185 | | | |
| *No (Ref.)* | | — | — | |
| *Yes* | | 0.66 | 0.58, 0.76 | <0.001 |
| **Parental monitoring (13y)** | 4,386 | | | |
| *Always/most of the time (Ref.)* | | — | — | |
| *Never/Hardly ever/Sometimes* | | 0.64 | 0.52, 0.79 | <0.001 |
| **Experienced any bullying (13y)** | 5,291 | | | |
| *No (Ref.)* | | — | — | |
| *Yes* | | 1.09 | 0.95, 1.25 | 0.209 |
| Abbreviations: OR = Odds Ratio, CI = Confidence Interval | | | | |

*Supplementary Table S3. Univariable associations between main analysis variables and being in the analytic sample (i.e. having data on parental smoking at age 12) using the full ALSPAC sample*

| Characteristic | N | OR | 95% CI | p-value |
|---|---|---|---|---|
| **Offspring substance use (13y)** | 6,687 | | | |
| *No (Ref.)* | | — | — | |
| *Yes* | | 0.71 | 0.62, 0.83 | <0.001 |
| **Offspring SMFQ scores (14y)** | 6,011 | | | |
| *<8 (Ref.)* | | — | — | |
| *≥8* | | 0.89 | 0.77, 1.03 | 0.112 |
| **Offspring current smoking (16y)** | 5,066 | | | |
| *No (Ref.)* | | — | — | |
| *Yes* | | 0.77 | 0.65, 0.91 | 0.002 |
| **Mother smoked in pregnancy (0y)** | 12,300 | | | |
| *No (Ref.)* | | — | — | |
| *Yes* | | 0.37 | 0.35, 0.41 | <0.001 |
| **Oldest parent's age at birth** | 8,320 | 1.05 | 1.05, 1.06 | <0.001 |
| **Lowest parental social class (0y)** | 11,514 | | | |
| *I/II (Ref.)* | | — | — | |
| *III* | | 0.67 | 0.61, 0.73 | <0.001 |
| *IV/V* | | 0.45 | 0.40, 0.50 | <0.001 |
| **Lowest parental education (0y)** | 11,768 | | | |
| *A level or above (Ref.)* | | — | — | |
| *O level* | | 0.65 | 0.59, 0.71 | <0.001 |
| *Vocational/CSE* | | 0.38 | 0.35, 0.42 | <0.001 |
| **Neighbourhood quality index (0y)** | 12,683 | 1.09 | 1.08, 1.11 | <0.001 |
| **Townsend deprivation score quintiles (8y)** | 7,519 | | | |
| *Q1 (Ref.)* | | — | — | |
| *Q2* | | 0.92 | 0.79, 1.08 | 0.316 |
| *Q3* | | 0.89 | 0.77, 1.03 | 0.122 |
| *Q4* | | 0.70 | 0.61, 0.81 | <0.001 |
| *Q5* | | 0.55 | 0.46, 0.66 | <0.001 |
| **Household income per week (8y)** | 7,080 | | | |
| *400+ (Ref.)* | | — | — | |
| *<400* | | 0.63 | 0.57, 0.70 | <0.001 |
| **Housing tenure (10y)** | 8,098 | | | |
| *Mortgaged/Owned (Ref.)* | | — | — | |
| *Rented/Other* | | 0.53 | 0.46, 0.60 | <0.001 |
| **Highest parental EPDS score (8y)** | 4,265 | 0.96 | 0.95, 0.97 | <0.001 |
| **Parental mental health condition (8y)** | 4,970 | | | |
| *No (Ref.)* | | — | — | |

| Characteristic | N | OR | 95% CI | p-value |
|---|---|---|---|---|
| *Yes* | | 0.60 | 0.52, 0.69 | <0.001 |
| **Sex assigned at birth** | 15,029 | | | |
| *Male (Ref.)* | | — | — | |
| *Female* | | 1.11 | 1.04, 1.18 | 0.002 |
| **Ethnicity** | 12,089 | | | |
| *White (Ref.)* | | — | — | |
| *Ethnically minoritised* | | 0.52 | 0.44, 0.62 | <0.001 |
| **Physical/emotional abuse (9y)** | 7,857 | | | |
| *No (Ref.)* | | — | — | |
| *Yes* | | 0.82 | 0.63, 1.08 | 0.148 |
| **Peer substance use (13y)** | 6,637 | | | |
| *No (Ref.)* | | — | — | |
| *Yes* | | 0.73 | 0.65, 0.82 | <0.001 |
| **Parental monitoring (13y)** | 5,670 | | | |
| *Always/most of the time (Ref.)* | | — | — | |
| *Never/Hardly ever/Sometimes* | | 0.81 | 0.69, 0.95 | 0.009 |
| **Experienced any bullying (13y)** | 6,764 | | | |
| *No (Ref.)* | | — | — | |
| *Yes* | | 0.86 | 0.77, 0.97 | 0.011 |
| Abbreviations: OR = Odds Ratio, CI = Confidence Interval | | | | |

*Supplementary Table S4. Multivariable association between outcome and missingness in the analytic sample where data is available for included variables (n = 2,105)*

| Characteristic | OR | 95% CI | p-value |
|---|---|---|---|
| **Offspring current smoking (16y)** | | | |
| *No (Ref.)* | — | — | |
| *Yes* | 0.80 | 0.63, 1.01 | 0.056 |
| **Parental daily smoking (12y)** | | | |
| *No (Ref.)* | — | — | |
| *Yes* | 1.00 | 0.78, 1.29 | 0.979 |
| **Offspring SMFQ scores (14y)** | | | |
| *<8 (Ref.)* | — | — | |
| *≥8* | 0.92 | 0.74, 1.13 | 0.416 |
| **Mother smoked in pregnancy (0y)** | | | |
| *No (Ref.)* | — | — | |
| *Yes* | 0.89 | 0.66, 1.20 | 0.444 |
| **Oldest parental age at birth** | 1.02 | 1.00, 1.03 | 0.062 |
| **Lowest parental education (0y)** | | | |
| *A level or above (Ref.)* | — | — | |
| *O level* | 0.75 | 0.60, 0.93 | 0.009 |
| *Vocational/CSE* | 0.70 | 0.54, 0.91 | 0.007 |
| **Lowest parental social class (0y)** | | | |
| *I/II (Ref.)* | — | — | |
| *III* | 1.00 | 0.81, 1.24 | 0.978 |
| *IV/V* | 0.70 | 0.51, 0.96 | 0.029 |
| **Neighbourhood quality index (0y)** | 1.02 | 0.98, 1.07 | 0.340 |

Abbreviations: OR = Odds Ratio, CI = Confidence Interval

*Supplementary Figure S4. Patterns of missing data across analysis variables in analytic sample (n = 6,741). The horizontal bars show the number of non-missing observations for each variable ("set size"), while the vertical bars represent the number of participants missing the same combination of variables ("intersection size"). Dots indicate which variables are jointly missing for each combination.*

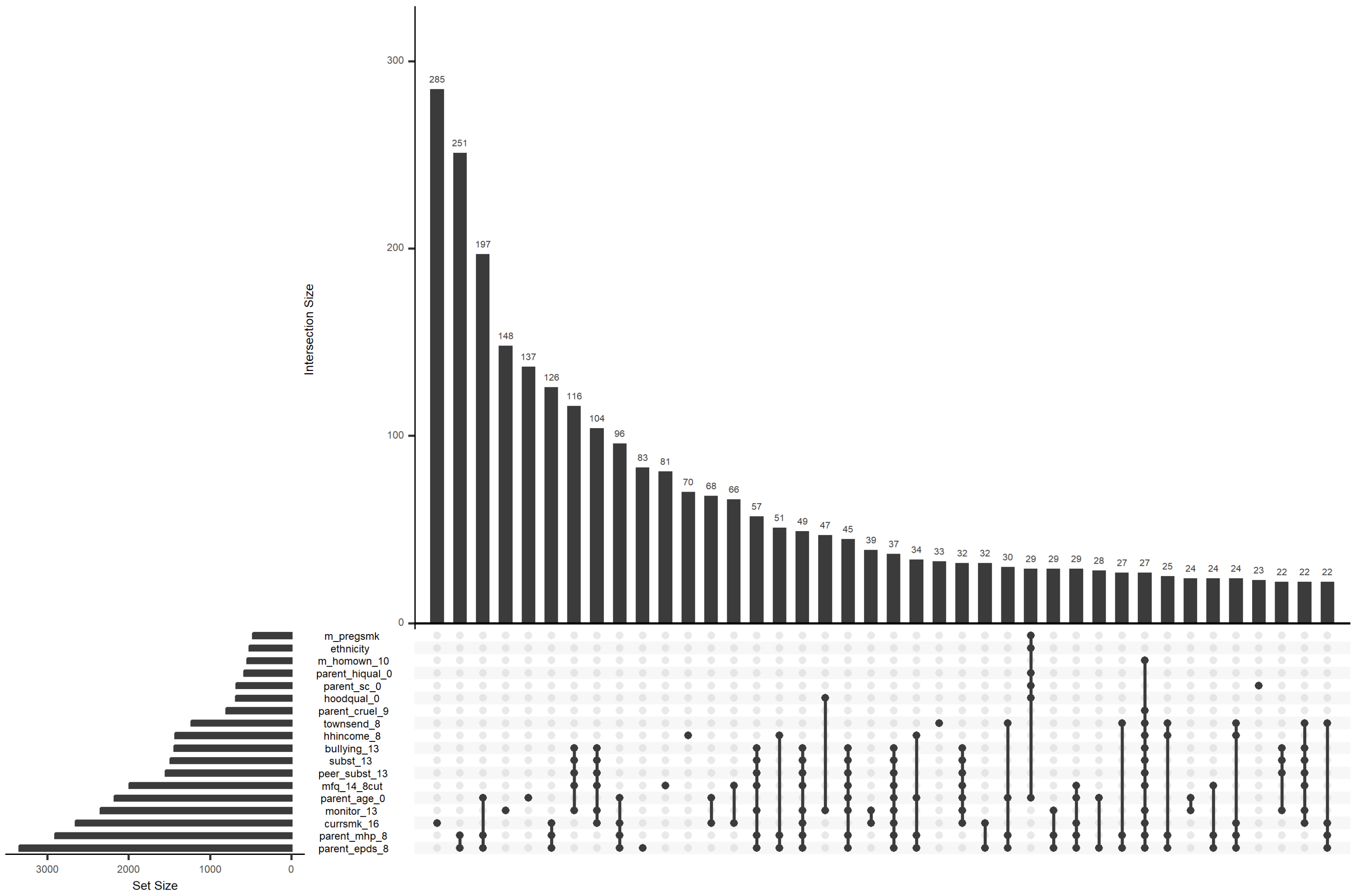

*Labels:* ***m_pregsmk*** *= Mother smoked in pregnancy (0y);* ***ethnicity*** *= Ethnicity;* ***m_homown_10*** *= Housing tenure (10y);* ***parent_hiqual_0*** *= Lowest parental education (0y);* ***parent_sc_0*** *= Lowest parental social class (0y);* ***hoodqual_0*** *= Neighbourhood quality index (0y);* ***parent_cruel_9*** *= Physical/emotional abuse (9y);* ***townsend_8*** *= Townsend deprivation score quintiles (8y);* ***hhincome_8*** *= Household income per week (8y);* ***bullying_13*** *= Experienced any bullying (13y);* ***subst_13*** *= Offspring substance use in past 6 months (13y);* ***peer_subst_13*** *= Peer substance use (13y);* ***mfq_14_8cut*** *= Offspring SMFQ scores; ≥8 (14y);* ***parent_age_0*** *= Oldest parental age at birth;* ***monitor_13*** *= Parental monitoring (13y);* ***currsmk_16*** *= Offspring current smoking (16y);* ***parent_mhp_8*** *= Parental mental health condition (8y);* ***parent_epds_8*** *= Highest parental EPDS score (8y)*

*Supplementary Table S5. G-computation formula estimates of the total and direct effects of parental smoking on offspring smoking and indirect effects via offspring depressive symptoms, using complete records (n = 1,050)*

| | Unadjusted for baseline & mediator-outcome confounders | | | Adjusted for baseline & mediator-outcome confounders | | |
|---|---|---|---|---|---|---|
| **Effect** | **OR** | **LCI** | **UCI** | **OR** | **LCI** | **UCI** |
| **No intermediate confounder** | | | | | | |
| **TCE** | 1.57 | 0.65 | 3.79 | 1.21 | 0.83 | 1.76 |
| **CDE** | 1.82 | 0.56 | 5.88 | 1.47 | 0.69 | 3.12 |
| **PNDE** | 1.58 | 0.64 | 3.89 | 1.25 | 0.81 | 1.95 |
| **TNIE** | 0.99 | 0.97 | 1.01 | 0.97 | 0.90 | 1.03 |
| **Including early offspring substance use (13y)** | | | | | | |
| **TCE** | 1.57 | 0.65 | 3.78 | 1.18 | 0.85 | 1.64 |
| **CDE** | 1.78 | 0.58 | 5.49 | 1.39 | 0.73 | 2.63 |
| **PNDE** | 1.60 | 0.64 | 3.99 | 1.22 | 0.82 | 1.81 |
| **TNIE** | 0.98 | 0.95 | 1.02 | 0.97 | 0.91 | 1.03 |

Abbreviations: OR = Odds Ratio, LCI = Lower Confidence Interval, UCI = Upper Confidence Interval, TCE = Total causal effect; CDE = Controlled direct effect; PNDE = Pure Natural Direct Effect; TNIE = Total Natural Indirect Effect

*Supplementary Table S6. Logistic regression estimates showing associations of parental smoking on early offspring substance use, using complete records (n = 1,050)*

| | Offspring substance use (13y) | | | | | |
|---|---|---|---|---|---|---|
| | Unadjusted[a] | | | Adjusted[a] | | |
| **Characteristic** | **OR** | **95% CI** | **p-value** | **OR** | **95% CI** | **p-value** |
| **Parental daily smoking (12y)** | | | | | | |
| *No (Ref.)* | — | — | | — | — | |
| *Yes* | 1.30 | 0.80, 2.06 | 0.274 | 1.02 | 0.53, 1.93 | 0.942 |

Abbreviations: OR = Odds Ratio, CI = Confidence Interval

[a] For baseline & mediator-outcome confounders

*Supplementary Table S7. Logistic regression estimates showing associations of parental smoking and early offspring substance use on offspring depressive symptoms, using complete records (n = 1,050)*

| | Offspring depressive symptoms (14y) | | | | | |
|---|---|---|---|---|---|---|
| | Unadjusted[a] | | | Adjusted[a] | | |
| **Characteristic** | **OR** | **95% CI** | **p-value** | **OR** | **95% CI** | **p-value** |
| **Parental daily smoking (12y)** | | | | | | |
| *No (Ref.)* | — | — | | — | — | |
| *Yes* | 1.58 | 1.09, 2.26 | 0.014 | 1.51 | 0.95, 2.38 | 0.076 |
| **Offspring substance use (13y)[b]** | | | | | | |
| *No (Ref.)* | — | — | | — | — | |
| *Yes* | 1.96 | 1.27, 3.00 | 0.002 | 1.22 | 0.72, 2.05 | 0.457 |

Abbreviations: OR = Odds Ratio, CI = Confidence Interval

[a] For baseline & mediator-outcome confounders

[b] Adjusted for parental smoking at offspring at age 12

*Supplementary Table S8. Logistic regression estimates showing associations of parental daily smoking, early offspring substance use and offspring depressive symptoms on later offspring smoking, using complete records (n = 1,050)*

| | Offspring smoking (16y) | | | | | |
|---|---|---|---|---|---|---|
| | Unadjusted[a] | | | Adjusted[a] | | |
| **Characteristic** | **OR** | **95% CI** | **p-value** | **OR** | **95% CI** | **p-value** |
| **Parental daily smoking (12y)** | | | | | | |
| *No (Ref.)* | — | — | | — | — | |
| *Yes* | 1.57 | 1.05, 2.32 | 0.026 | 1.22 | 0.74, 1.99 | 0.426 |
| **Offspring substance use (13y)[b]** | | | | | | |
| *No (Ref.)* | — | — | | — | — | |
| *Yes* | 5.85 | 3.84, 8.89 | <0.001 | 4.58 | 2.76, 7.64 | <0.001 |
| **Offspring SMFQ scores (14y)[b]** | | | | | | |
| *<8 (Ref.)* | — | — | | — | — | |
| ≥8 | 1.46 | 0.98, 2.14 | 0.056 | 1.03 | 0.66, 1.58 | 0.892 |
| **Offspring SMFQ scores (14y)[b,c]** | | | | | | |
| *<8 (Ref.)* | — | — | | — | — | |
| ≥8 | 1.26 | 0.83, 1.88 | 0.274 | 1.00 | 0.63, 1.55 | 0.988 |

Abbreviations: OR = Odds Ratio, CI = Confidence Interval

[a] For baseline & mediator-outcome confounders

[b] Adjusted for parental smoking at offspring at age 12

[c] Adjusted for offspring substance use at age 13

*Supplementary Table S9. Unadjusted and adjusted interaction term between offspring depressive symptoms at age 14 and parental daily smoking at age 12 on current smoking at age 16 based on complete records analysis (n = 1,050) and multiple imputation (n = 6,729)*

| | Unadjusted for baseline & mediator-outcome confounders | | | Adjusted for baseline & mediator-outcome confounders | | |
|---|---|---|---|---|---|---|
| **Characteristic** | **OR** | **95% CI** | **p-value** | **OR** | **95% CI** | **p-value** |
| *Complete records estimate* | | | | | | |
| **Parental daily smoking (12y)** | | | | | | |
| *No (Ref.)* | — | — | | — | — | |
| *Yes* | 1.82 | 1.13, 2.85 | 0.011 | 1.52 | 0.87, 2.60 | 0.134 |
| **Offspring SMFQ scores (14y)** | | | | | | |
| *<8 (Ref.)* | — | — | | — | — | |
| *≥8* | 1.72 | 1.09, 2.65 | 0.016 | 1.26 | 0.77, 2.03 | 0.343 |
| **Parental daily smoking (12y) * Offspring SMFQ scores (14y)** | | | | | | |
| *Yes * ≥8* | 0.52 | 0.20, 1.27 | 0.161 | 0.43 | 0.16, 1.12 | 0.092 |
| *Multiple imputation estimate* | | | | | | |
| **Parental daily smoking (12y)** | | | | | | |
| *No (Ref.)* | — | — | | — | — | |
| *Yes* | 2.13 | 1.74, 2.61 | <0.001 | 1.51 | 1.19, 1.91 | <0.001 |
| **Offspring SMFQ scores (14y)** | | | | | | |
| *<8 (Ref.)* | — | — | | — | — | |
| *≥8* | 1.83 | 1.43, 2.35 | <0.001 | 1.37 | 1.05, 1.79 | 0.019 |
| **Parental daily smoking (12y) * Offspring SMFQ scores (14y)** | | | | | | |
| *Yes * ≥8* | 0.96 | 0.67, 1.39 | 0.844 | 0.94 | 0.65, 1.38 | 0.758 |
| Abbreviations: OR = Odds Ratio, CI = Confidence Interval | | | | | | |
